# Deciphering how early life adiposity influences breast cancer risk using Mendelian randomization

**DOI:** 10.1101/2021.06.22.21259310

**Authors:** Marina Vabistsevits, George Davey Smith, Eleanor Sanderson, Tom G Richardson, Bethan Lloyd-Lewis, Rebecca C Richmond

**Author notes:** Corresponding author: Marina Vabistsevits, MRC Integrative Epidemiology Unit, Population Health Sciences, Bristol Medical School, University of Bristol, Oakfield House, Oakfield Grove, Bristol BS8 2BN, UK.

## Abstract

Studies suggest that adiposity in childhood may reduce the risk of breast cancer in later life. The biological mechanism underlying this effect is unclear but is likely to be independent of body size in adulthood. Using a Mendelian randomization framework, we investigated 18 hypothesised mediators of the protective effect of childhood adiposity on later-life breast cancer, including hormonal, reproductive, physical, and glycaemic traits.

Our results indicate that, while most of the hypothesised mediators are affected by childhood body size, only IGF-1, testosterone, age at menarche and age at menopause influenced breast cancer risk. However, accounting for those traits in multivariable Mendelian randomization showed that the protective effect of childhood body size still remained. This suggests either a direct effect of childhood body size on breast cancer risk or mediation via other pathways not considered.

Our work presents a framework for the systematic exploration of potential biological mediators of disease in Mendelian randomization analysis.

## Introduction

Breast cancer is the leading cause of cancer-related deaths among women, with 1 in 8 at risk of developing the disease in high-income countries [1], [2]. Although breast cancer mortality has declined over recent decades, mostly due to improved treatments and personalised diagnoses [3], the incidence of the disease has been steadily increasing by 3.1% annually since the 1980s [4]. Reducing the incidence rates will depend on better understanding and communicating modifiable risk factors both population-wide and targeted at women with an increased risk [5].

The second-largest preventable risk to all cancers after smoking is obesity [6], which has been extensively studied in relation to breast cancer. In observational studies, there is consistent evidence for a positive association of increased body mass index (BMI) with post-menopausal breast cancer, but an inverse association with pre-menopausal breast cancer [5], [6]. This may be explained by varying levels of exposure to oestrogen in overweight compared with normal-weight women. Pre-menopause, overweight women have longer anovulatory cycles, thereby decreasing their exposure to ovarian hormones which has been suggested to reduce their breast cancer risk [9]. Post-menopause, adipose tissue is the main source of oestrogen biosynthesis [10]. This increases exposure to oestrogen in overweight compared to normal-weight women, which may explain their higher risk of breast cancer after menopause.

While previous conclusions have largely been drawn from observational epidemiological studies [11]–[13], several Mendelian randomization (MR) analyses have shown a contrary result, where genetically-instrumented BMI is inversely associated with the risk of both pre- and post-menopausal breast cancer [14], [15]. Since the genetic variants used to instrument BMI are set at birth, they should not be affected by environmental factors in later life. Thus, Guo *et al* [14] hypothesised that the positive association between high BMI and post-menopausal breast cancer risk seen observationally may reflect adiposity and weight gain later in life. This is supported by findings of an inverse association between early-life (childhood and adolescence) BMI with breast cancer risk [11], [12]. A recent MR study [16] using genetic variants related to childhood body size showed the same protective effect on breast cancer risk. Furthermore, using a multivariable MR analysis that accounted for adult body size, this study indicated that the protective effect of childhood body size influences breast cancer risk directly, independently of adult body size. However, the mechanism by which larger body size during childhood may reduce future breast cancer risk is not understood. Deciphering the mediating pathway between early-life adiposity and breast cancer would be of great interest for identifying targets of intervention, since advocating weight gain in childhood is not recommended.

Many established breast cancer risk factors are plausible candidate mediators of the protective effect of early-life body size, since they have been found to be influenced or associated with childhood adiposity (Figure 1). For example, body fatness during childhood may protect against breast cancer risk through hormonal pathways (e.g. IGF-1, oestradiol, testosterone, SHBG [17], [18]). It has also been shown that incidence of breast cancer, particularly oestrogen receptor-positive (ER+) breast cancer, is substantially driven by changes in reproductive patterns, including parity and age at menarche, first birth and menopause, [19], [20]), events that are also influenced by increased adiposity in childhood [21], [22]. Physical traits such as breast mammographic density (MD), which is an established risk factor of breast cancer [23], are also affected by body fatness in early life [24], [25]. Finally, glycaemic traits have been extensively studied in relation to BMI and, while they have been inconsistently associated with breast cancer [26]–[28], are also of interest as potential intermediates.

**Figure 1.**
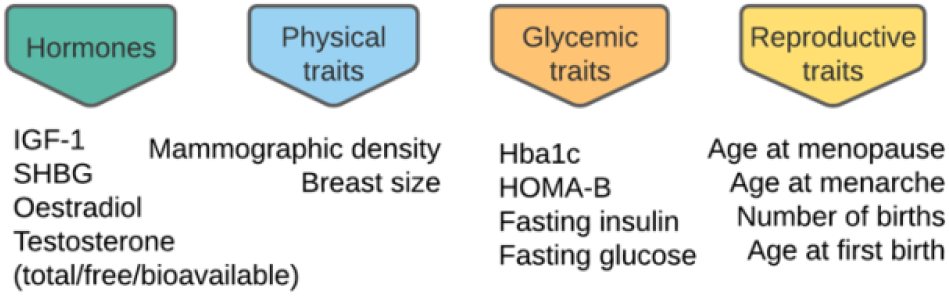
Trait categories that may be influenced by childhood adiposity to affect breast cancer.

In this work, we aimed to decipher the link between increased childhood body size and breast cancer risk by assessing a variety of potential mediators of this effect within a Mendelian randomization framework [29], [30]. We characterised the effects of four groups of mediators that may be influenced by childhood body size to affect breast cancer risk: sex hormones, reproductive traits, glycaemic traits, and physical traits. This was done using several extensions of the basic MR principle, including two-sample MR [31], two-step MR [32], and multivariable MR [28], with results then integrated within a mediation framework.

## Results

### Study overview

Using genome-wide association study (GWAS) summary statistics for childhood body size [16], breast cancer [51] and 18 reproductive [36], [38], [39], glycaemic [40]–[45], physical [15], [48]–[50] and hormonal traits, we set up an MR framework to investigate the role of these traits as plausible mediators of the childhood body size effect on breast cancer risk in later life. Firstly, we performed two-step MR to identify traits influenced by childhood body size that have a causal effect on breast cancer (Figure 2). Subsequently, traits were assessed in a multivariable MR analysis to assess the magnitude of their direct effect on breast cancer risk, followed by a mediation analysis to quantify the indirect effect. The main exposure and outcome datasets used in the analysis comprised 246K female participants from UK Biobank (childhood body size) and 228K participants from Breast Cancer Association Consortium, BCAC, (breast cancer cases and controls) respectively. An overview of the GWAS datasets used as mediators is presented in Table 1.

**Figure 2.**
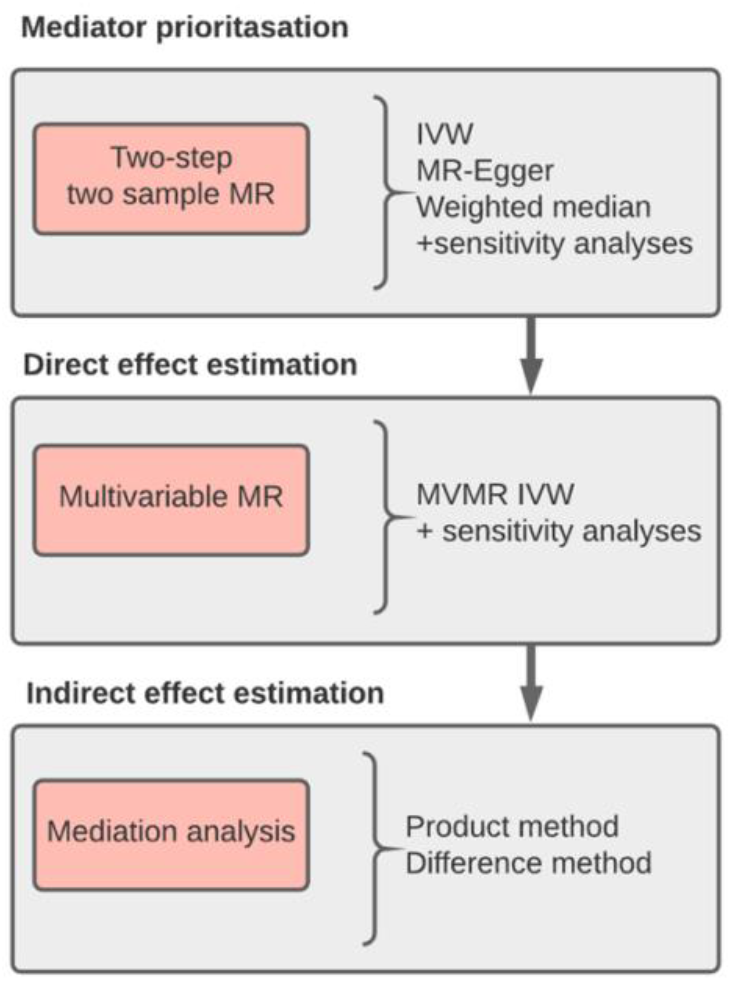
Study design. MR – Mendelian randomization

**Table 1.** Summary of GWAS datasets used as mediators in the study. *Mediator trait:* name of the mediator as used throughout the paper; if there were several mediator GWAS available, **\*** indicates the one that was used in the main analysis/plots. *Study:* cohort/publication/author of the dataset. *Data source*: how data was created or acquired; IEU GWAS pipeline is the MRC IEU in-house GWAS pipeline for processing UK Biobank data [35]; OpenGWAS [36] (gwas.mrcieu.ac.uk) is the MRC IEU-based platform that hosts summary statistics of publicly available GWAS studies, the datasets IDs are provided; MAGIC ftp server is available here: www.magicinvestigators.org/downloads/; Supplementary data refers to the data available with the publication in the ‘Study’ column; GWAS catalog [37] hosts top hits from GWAS studies. *Sample size:* number of samples in each GWAS. *GWAS size:* number of SNPs used to run the GWAS. *Sample sex:* GWAS female/male composition. *SNPs at 5e-08:* number of genome significant SNPs (top hits). *Units:* most are in standard deviation (SD) unless otherwise specified. *PMID:* PubMed ID.

| Mediator trait | Study | Data source | Sample size | GWAS size | Sample sex | SNPs at 5e-08 | Units | PMID |
| --- | --- | --- | --- | --- | --- | --- | --- | --- |
| *Hormones* |  |  |  |  |  |  |  |  |
| IGF-1 | UK Biobank | IEU GWAS pipeline | 246,284 | 12,321,875 | F | 375 | SD | this study |
| Oestradiol | UK Biobank | IEU GWAS pipeline | 53,491 | 12,321,875 | F | 1 | SD | this study |
| SHBG | UK Biobank | IEU GWAS pipeline | 222,491 | 12,321,875 | F | 199 | SD | this study |
| Testosterone (bioavailable) | UK Biobank | IEU GWAS pipeline | 186,700 | 12,321,875 | F | 124 | SD | this study |
| Testosterone (free) | UK Biobank | IEU GWAS pipeline | 186,700 | 12,321,875 | F | 129 | SD | this study |
| Testosterone (total) | UK Biobank | IEU GWAS pipeline | 206,604 | 12,321,875 | F | 115 | SD | this study |
| *Reproductive traits* |  |  |  |  |  |  |  |  |
| Age at first live birth | UK Biobank [36] | OpenGWAS: ukb-b-12405 | 170,498 | 9,851,867 | F | 35 | SD | n/a |
| Age at menarche * | Perry JR 2014 [38] | OpenGWAS: ieu-a-1095 | 182,416 | 2,441,816 | F | 68 | SD | 25231870 |
| Age at menarche | UK Biobank [36] | OpenGWAS: ukb-b-17422 | 143,819 | 9,851,867 | F | 114 | SD | n/a |
| Age at menopause * | Day 2015 [39] | OpenGWAS: ieu-a-1004 | 69,360 | 2,418,696 | F | 42 | SD | 26414677 |
| Age at menopause | UK Biobank [36] | OpenGWAS: ukb-b-3768 | 243,944 | 9,851,867 | F | 200 | SD | n/a |
| Number of births | UK Biobank [36] | OpenGWAS: ukb-b-1209 | 250,782 | 9,851,867 | F | 11 | SD | n/a |
| *Glycaemic traits* |  |  |  |  |  |  |  |  |
| Fasting glucose * | Lagou 2021 [40] | MAGIC ftp | 73,089 | 8.7M | F | 22 | SD | 33402679 |
| Fasting glucose | Manning AK 2012 [41] | OpenGWAS: ieu-b-113 | 58,074 | 2,628,880 | M/F | 22 | SD | 22581228 |
| Fasting glucose | Scott 2012 [42] | OpenGWAS: ieu-b-114 | 133,010 | 64,494 | M/F | 35 | SD | 22885924 |
| Fasting insulin * | Lagou 2021 [40] | MAGIC ftp | 50,404 | 8.7M | F | 4 | SD | 33402679 |
| Fasting insulin | Manning AK 2012 [41] | OpenGWAS: ieu-b-115 | 51,750 | 2,627,849 | M/F | 4 | SD | 22581228 |
| Fasting insulin | Scott 2012 [42] | OpenGWAS: ieu-b-116 | 108,557 | 64,484 | M/F | 13 | SD | 22885924 |
| HBa1c * | Wheeler 2017 [43] | MAGIC ftp | 123,665 | 9M | M/F | 37 | SD | 28898252 |
| HBa1c | Soranzo 2010 [44] | OpenGWAS: ieu-b-103 | 46,368 | 2,576,680 | M/F | 11 | SD | 20858683 |
| HOMA-B | Dupuis J 2011 [45] | OpenGWAS: ieu-b-117 | 46,186 | 2,456,946 | M/F | 4 | SD | 20081858 |
| *Physical traits* |  |  |  |  |  |  |  |  |
| Breast size * | 23andMe/ Nick Sern Ooi 2019 [15] | supplementary material | 33,790 | 13M | F | 7 | cup size | 31243447 |
| Breast size | 23andMe/ Pickrell2016 [46] | GWAS catalog/ supplementary material | 33,790 | 10M | F | 10 | unit | 27182965 |
| Breast size | 23andMe/ Eriksson2012 [47] | GWAS catalog/ supplementary material | 16,175 | 10M | F | 6 | cup size | 22747683 |
| Mammographic density * (dense/non/%) | Sieh 2020 [48] | supplementary material | 24,192 | 650K | F | 15/10/9 | SD | 33037222 |
| Mammographic density (dense/non/%) | Lindström 2014 [49] | GWAS catalog | ~14,788 | 2.5M | F | 7/1/3 | unit/ % | 25342443 |
| Mammographic density (dense/non/%) | Brand 2018 [50] | supplementary material | 9,498 | 500K | F | 5/2/3 | volume (cm3)/% | 29665850 |

### Two-step Mendelian randomization

For each mediator, we conducted two-step MR, where each step is an independent univariable two-sample MR analysis. In step 1, childhood body size was used as the exposure and a mediator trait as the outcome; in step 2, the mediator was used as the exposure and breast cancer as the outcome (Figure 3B). This analysis provided insights into the presence of a causal effect on/from the mediators and was used as a mediator prioritisation step. The results obtained using the IVW (inverse-variance weighted) method are presented in Figure 4.

**Figure 3.**
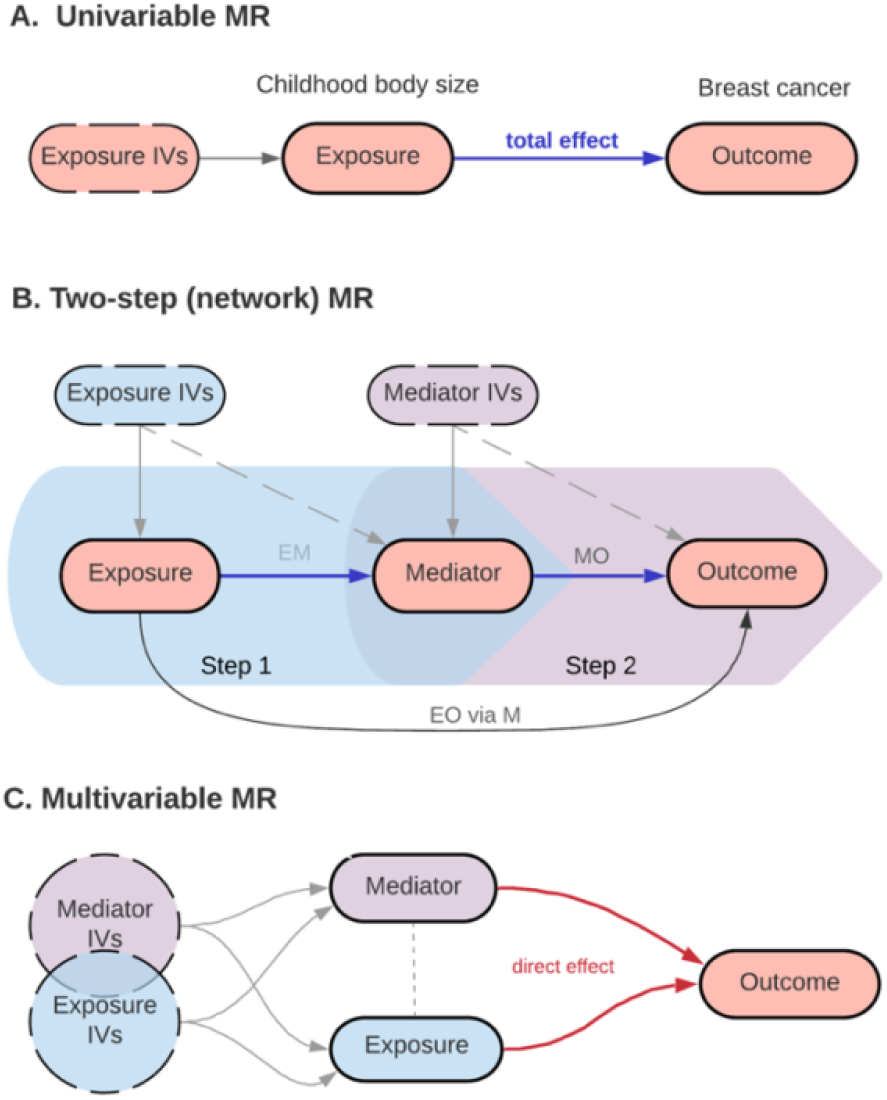
Mendelian randomization (MR) analysis methods used in the study. *(A) Univariable MR:* simple two-sample setup, measuring the total effect of exposure (childhood body size) on the outcome (breast cancer). *(B) Two-step (network) MR:* two sets of two-sample MR – step1 (EM): the total effect of exposure on the mediator, step 2 (MO): the total effect of the mediator on the outcome, allowing the measurement of the indirect effect of exposure on outcome via mediator (EO via M). *(C) Multivariable MR:* both exposure and mediator are accounted for in a single model; the direct effect of both is estimated. **IVs** – instrumental variables; **EM** – exposure-mediator; **MO** – mediator-outcome; **EO** – exposure-outcome.

**Figure 4.**
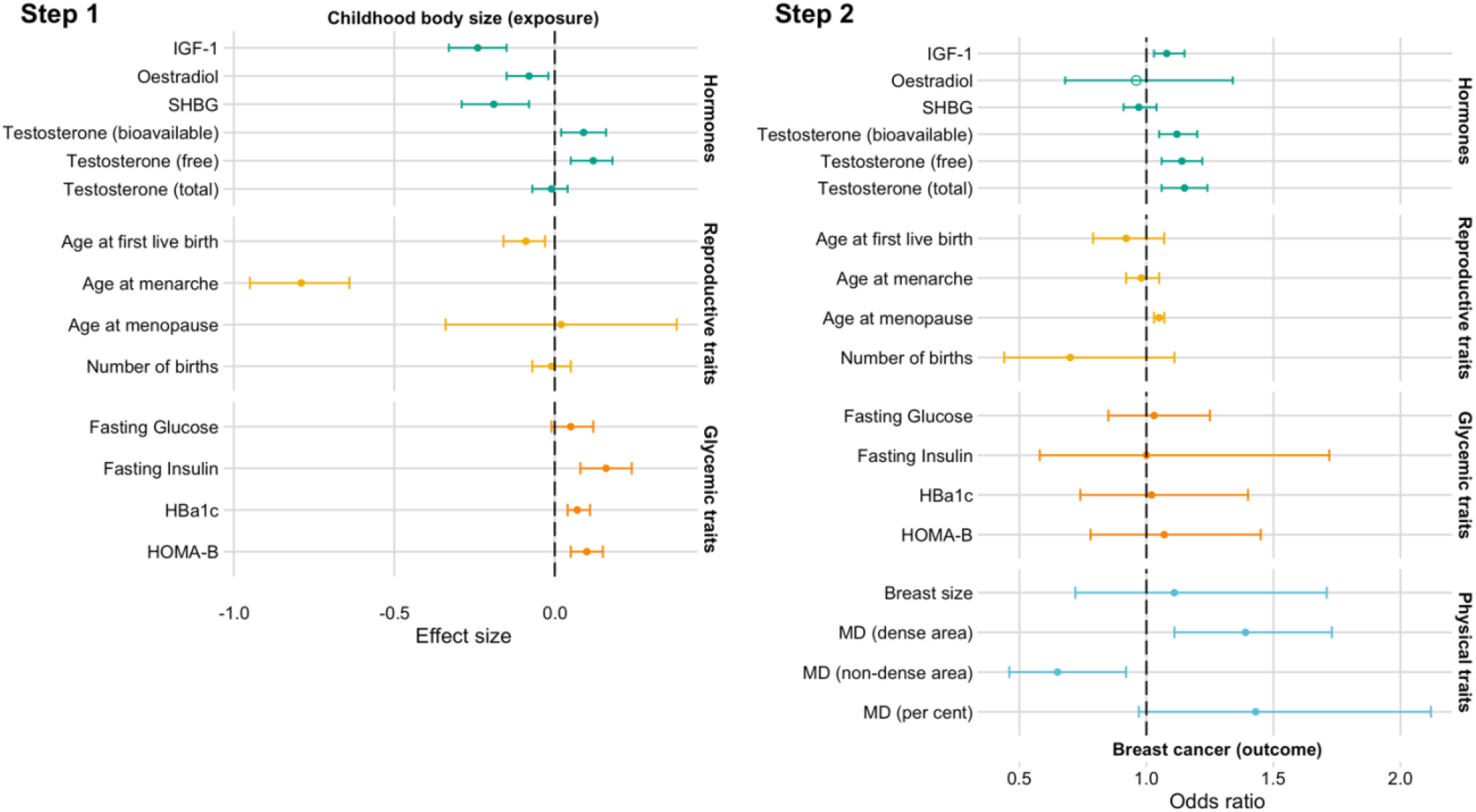
Two-step Mendelian randomization results evaluating four groups of potential mediators (hormones, reproductive traits, glycaemic traits, physical traits). **Step 1**: Plots showing the effect of childhood body size on the mediators (univariable MR). The effect is measured as the standard deviation (SD) change in mediator per body size category change. **Step 2**: Plots showing the odds of breast cancer per SD higher mediators (unless otherwise specified in Table 1; univariable MR). Bars indicate 95% confidence intervals around the point estimates from IVW analyses (in step 1: effect size/beta, in step 2: odds ratio), except oestradiol (the estimate is based on a single Wald ratio and is indicated by an empty circle shape). MD – mammographic density. The presented data is available in Supplementary Tables S2 and S3. The details about the datasets are provided in Table 1.

Among the hormones investigated, evidence of an effect in both MR steps was observed for IGF-1 and free and bioavailable testosterone (Figure 4). Increased childhood body size was associated with a reduction in IGF-1 (effect size per standard deviation, -0.24, 95% confidence interval: [-0.33: -0.15]), while IGF-1 had a positive effect on breast cancer risk (odds ratio per standard deviation, OR, 1.08 [1.03: 1.15]). Bioavailable and free testosterone estimates were similarly affected by childhood body size (0.09 [0.02: 0.16] and 0.12 [0.05: 0.18], respectively), and also had a similar positive effect on breast cancer risk (OR: 1.12 [1.05: 1.2] and OR: 1.14 [1.06: 1.22], respectively). Total testosterone had a positive effect on breast cancer (OR: 1.15 [1.06: 1.24], however, there was little evidence to suggest it was affected by childhood body size (−0.01 [-0.07: 0.04]). Oestradiol (−0.08 [-0.15: -0.02]) and SHBG (−0.19 [-0.29: -0.08]) were both inversely affected by increased childhood body size, but there was limited evidence of them affecting the risk of breast cancer (OR: 0.96 [0.68: 1.34] and OR: 0.97 [0.91: 1.04], respectively).

Analysis of reproductive traits showed the greatest effect of high childhood body size on age at menarche (−0.79 [-0.95: -0.64], effect size per standard deviation, 95% CIs), as well as an effect on age at first birth (−0.09 [-0.16: -0.03]). There was little evidence that age at menopause and number of births were affected by increased childhood body size (0.02 [-0.34: 0.38] and -0.01 [-0.07: 0.05], respectively). Age at menopause was found to have a positive effect on breast cancer (OR: 1.05 [1.03: 1.07]). The OR point estimates of other reproductive traits were inverse, but overall, there was little evidence of their effect on breast cancer (age at menarche – OR: 0.98 [0.92: 1.05], age at first birth – OR: 0.92 [0.79: 1.07], number of births – OR: 0.7 [0.44: 1.11]). The estimates for age at menarche and age at menopause from the other data source were in agreement with the results in Figure 4 (Supplementary Tables S2 and S3).

The tested glycaemic traits generally had strong evidence of being positively affected by increased childhood body size, although there were some inconsistent results using different data sources for the same traits (e.g. fasting insulin, see Supplementary Table S2). The estimated effects were – fasting insulin: 0.16 [0.08: 0.24], fasting glucose: 0.05 [-0.01: 0.12], Hba1c: 0.07 [0.04: 0.11], HOMA-B: 0.1 [0.05: 0.15]. However, none of the glycaemic traits had a substantial effect on breast cancer risk (fasting insulin OR: 1.00 [0.58:1.72], fasting glucose OR: 1.03 [0.85: 1.25], Hba1c OR: 1.02 [0.74: 1.4], HOMA-B OR: 1.06 [0.78: 1.45].

For the physical traits, we were only able to perform the second step of the MR analysis (i.e. the effect of the trait on breast cancer) since we did not have access to the full GWAS summary data. There was limited evidence for an effect from breast size on breast cancer risk (OR: 1.11 [0.72: 1.71]), as previously shown by Nick Sern Ooi *et al*. (2019) using the same data. Among the mammographic density (MD) phenotypes, the dense area of the breast had a positive effect (OR: 1.39 [1.11: 1.73]) and non-dense area had a negative effect (OR: 0.65 [0.46: 0.92]) on breast cancer risk. There was also a positive effect of per cent MD, although with wider confidence intervals (OR: 1.43 [0.97: 2.12]).

We also repeated the second step MR analyses using breast cancer data stratified into oestrogen receptor-positive (ER+) and negative (ER-) groups [51] (Supplementary Tables S9 and S10). Overall, similar findings were identified for the ER+ cases across all mediators, with stronger effects for MD phenotypes. None of the investigated hormones that had an effect on overall breast cancer risk showed evidence of an effect on ER-cases. Among the reproductive traits, the direction of effect switched from inverse to positive for age at menarche, whereas the estimate for age at menopause shifted closer to the null for ER-cases. No strong effects of the glycaemic traits were found on the ER-cases, consistent with ER+ and total breast cancer sample.

### Multivariable Mendelian randomization

We next performed MVMR analyses with childhood body size and each mediator in turn, in relation to breast cancer risk. This allowed us to establish the direct effect of childhood body size on breast cancer risk after accounting for each mediator (Figure 3C). MVMR was performed only on those mediators that showed the evidence of effect in at least one of the two-step MR steps and where the sensitivity analyses of both steps showed consistent results (see **Sensitivity analysis** and prioritisation logic in Supplementary Materials, Section A).

#### Direct effects of childhood body size

The direct effect of childhood body size on breast cancer after accounting for each mediator estimated using the IVW-MVMR method are presented in Figure 5 (values in Supplementary Table S4). Compared with the total effect of childhood body size on breast cancer from univariable MR, which had an of OR 0.66 [0.57, 0.76]), the direct effects (ORs) of childhood body size varied between 0.59 (age at menarche) and 0.67 (total testosterone). This indicates no substantial change in the effect of childhood body size on breast cancer after accounting for each of the potential mediators, with effects consistent between overall, ER+ and ER-breast cancer (Supplementary Tables S11-S12).

**Figure 5.**
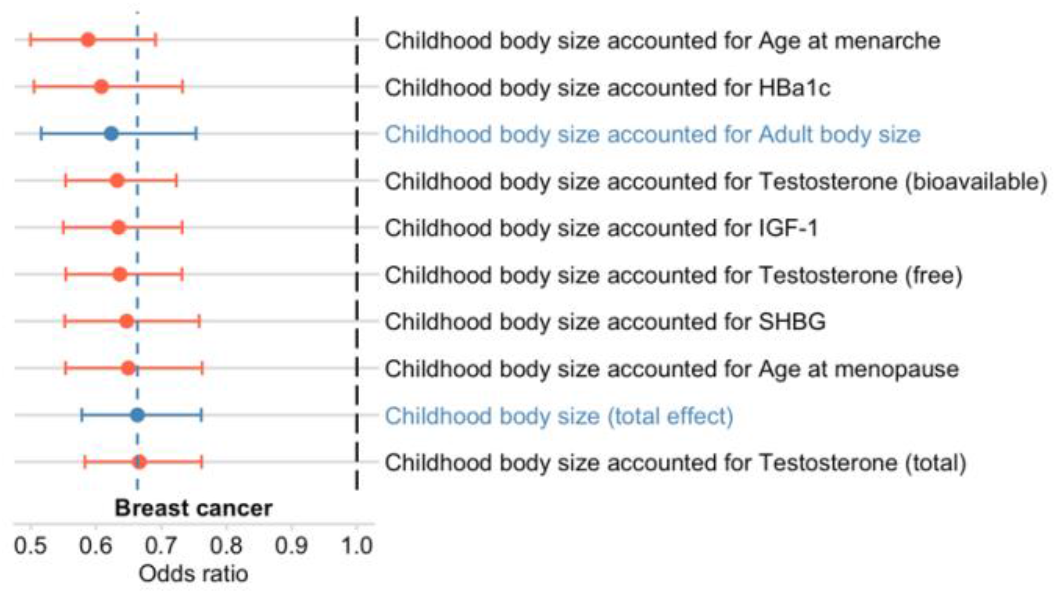
Multivariable Mendelian randomization results of childhood body size direct estimates accounted for selected mediators. The plot shows the odds of breast cancer per SD change in the direct effect of childhood body size accounted for mediators (red). The estimates in blue are total (OR: 0.66 [0.57: 0.76]) and direct (i.e. accounted for adult body size, OR: 0.62 [0.51: 0.75]) effect estimates of childhood body size on breast cancer risk were re-calculated from the original study [15] (using the thresholds defined in the Methods to match other MR analysis in this study), included here for comparison. The vertical blue line shows the value of total childhood body size effect relative to the direct estimates. Bars indicate 95% confidence intervals around the point estimates from IVW-MVMR. The estimated values and the numbers of genetic variants included in the analyses are given in Supplementary Table S4. The details about the datasets are given in Table 1.

We also performed analyses with adult body size and childhood height as third exposures in MVMR (in addition to childhood body size and the mediator) in two separate analyses. Again, no substantial change in the direct effect of childhood body size was not observed in the presence of the additional exposures (Supplementary Tables S13 and S14).

#### Direct effects of mediators

Overall, the direct effects of mediators on breast cancer risk were similar to their total effects once accounted for childhood body size (Supplementary Figure S2, values in Supplementary Table S4). Among the hormones investigated, the direction and size of effect remained the same (+/-0.01 OR) for all measures (IGF-1, SHBG, and the three measures of testosterone). Among the reproductive traits, MVMR indicated a protective direct effect of increasing age at menarche once accounted for childhood body size (OR: 0.92 [0.86: 0.99]), and the effect was even stronger using the alternative age at menarche GWAS source (OR: 0.82 [0.74: 0.91], Supplementary Table S4). The positive effect of age at menopause remained consistent with univariable MR results (OR: 1.05 [1.03: 1.07]). The point estimates of Hba1c shifted from the null, but wide confidence intervals still indicate little evidence of the effect (OR: 1.06 [0.83:1.35]). Overall, similar findings to overall breast cancer were identified for the ER+ cases across all mediators. However, in the ER-cases, similar to trends observed in step 2 in two-step MR, there was limited evidence for an effect of the mediators, with the exception of a protective effect of age at first birth in relation to ER-cases (OR: 0.72 [0.56: 0.94] compared with ER+ OR: 0.96 [0.79: 1.17]) (Supplementary Tables S11 and S12).

In the MVMR analyses also accounting for adult body size effect, the mediator estimates were not significantly affected. However, accounting for childhood height reduced the direct estimates for most mediators and attenuated IGF-1 and age at menarche effects to overlap the null (Figure S3 in Supplementary Materials Section C and Tables S13 and S14).

### Sensitivity analysis

To investigate the potential violations of the MR assumptions and validate the robustness of the two-sample MR results from the IVW approach, we performed additional MR analyses using MR-Egger [52] and weighted median [53] approaches, both of which are more robust to pleiotropy. The Egger intercept was used to formally test for the presence of horizontal pleiotropy, and Cochran’s Q statistic was calculated to quantify the extent of heterogeneity among SNPs. The sensitivity analyses of MVMR included tests for instrument strength and horizontal pleiotropy. Performance in the sensitivity tests was used as a selection tool for mediator inclusion in the downstream analyses (Supplementary Materials, Section A).

The estimated effects for hormones and reproductive traits were consistent across sensitivity analyses. The results obtained for some of the glycaemic traits, however, were variable and should be interpreted with caution. For all pairs of childhood body size and a mediator which were prioritised for MVMR analysis, conditional F-statistics were > 10, indicating that weak instrument bias is unlikely to be present. The presence of directional pleiotropy was assessed by estimating Q_A_ statistics, which consistently indicated excess heterogeneity and so the potential for pleiotropy. The sensitivity analysis details for all mediator groups are available in Supplementary Materials (Section B), and Tables S6-S8.

Lastly, the violation of two-sample MR requirement of having two non-overlapping datasets for exposure and outcome traits, i.e. “winner’s curse”, which was present for hormone traits (step 1 of two-step MR), was addressed with the ‘split-sample’ approach (see **Methods**). Results of this analysis were consistent with the full sample analysis (Supplementary Materials Section D and Tables S15 and S16).

### Mediation analysis

Next, we carried out mediation analysis to estimate the indirect effect of childhood body size on breast cancer risk via selected mediators, using the effect estimates from two-step MR, MVMR, and the total effect (Figure 3A, Supplementary Table S1). This analysis was restricted to mediators that showed evidence of an effect in MVMR and that had substantial instrument strength (Supplementary Materials, Section A).

Using a simulation analysis to compare available mediation methods (Supplementary Materials, Section E), the Product method was chosen with reasonably high confidence as the main mediation analysis approach, with the Delta method as the corresponding SE/CI estimation technique. The indirect effect results are displayed in Figure 6 and the estimates are available in Supplementary Table S17.

**Figure 6.**
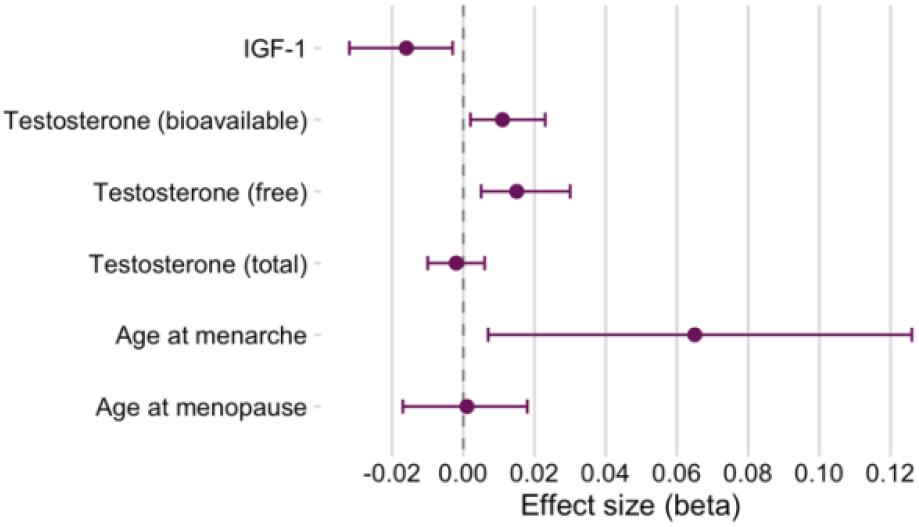
Mediation analysis results: indirect effect of childhood body size via a mediator. The indirect effect was estimated as the product of coefficients of the total effect of exposure on the mediator (step 1 of two-step MR) and the direct effect of a mediator on the outcome (MVMR), i.e. Product method, and the CIs were calculated based on SE estimated using Delta method.

The indirect effects were modest, although there was some evidence for an inverse indirect effect via IGF-1 (−0.016 [-0.032: -0.003]), suggesting potential mediation via this trait. The estimated proportion of the mediated effect via IGF-1 was 0.039, indicating that any potential mediation via IGF-1 would only account for 3.9% of the total effect. Conversely, the alternative mediation approach (Difference method) estimated the IGF-1 indirect effect to be positive (described in Supplementary Materials, Section E). Thus, the observed negative effect of IGF-1 requires further investigation. Mediation analysis also showed a positive indirect effect of childhood body size via age at menarche (0.065 [0.007: 0.126]) and via free and bioavailable testosterone (0.015 [0.005: 0.03] and 0.011 [0.002: 0.023] respectively, in contrast to the negative total effect of childhood body size on breast cancer.

## Discussion

Previous observational and MR studies indicate that early life body size has a protective effect on risk of breast cancer [11], [12], [16]. In this study, we investigated whether a number of breast cancer risk factors served as potential mediators of this protective effect using large genome-wide association datasets and a series of Mendelian randomization methods. Using two-step MR, we identified IGF-1, SHBG, testosterone, Hb1ac, age at menarche and age at menopause as plausible mediators based on the effect of childhood body size on these traits, and their effect on breast cancer risk. However, when applied in a multivariable MR framework, none of these traits appeared to substantially mediate the protective effect of early life body size on breast cancer risk.

The results of our MR analysis of selected hormones on breast cancer are supported by recent MR studies, with similar effects observed for IGF-1 in Murphy *et al* (2020) [54] and SHBG and testosterone in Ruth *et al* (2020) [55]. Conversely, the negative association of SHBG with breast cancer risk adjusted for BMI observed in Dimou *et al* (2019) [56] had limited evidence of effect in our study, likely due to sex-specific analysis and differences in sample size. The effects observed for IGF-1 in two-step MR is in agreement with observational studies [17], [57]. IGF-1 is known to play an important role in breast tissue differentiation and mammary gland function [58], and during normal development, levels of IGF-1 gradually rise from birth to puberty, followed by a decrease in response to growth hormones [59]. The finding of lower adult IGF-1 in response to larger body size during childhood may be a part of the mechanism through which early life adiposity influences breast cancer risk. Our mediation analysis showed that childhood body size may have a negative effect on breast cancer indirectly via IGF-1. However, this result requires further investigation as the estimated indirect effect was relatively small (accounting for 3.9% of the total effect), and inconsistent across mediation methods. While there was some evidence for indirect effects of childhood body size via age at menarche, as previously reported in [16], [60], and testosterone levels, this was in the positive direction in opposition to the total inverse effect of childhood body size on breast cancer. Lastly, it was not feasible to confidently assess the effect of oestradiol on breast cancer in MR analyses due to the limited number of genetic instruments in the available data, but as oestradiol has been observationally associated with breast cancer [18], its mediating role remains of interest.

We also reviewed the effects in ER- and ER+ cancer samples, which can proxy for younger/older or pre-/post-menopausal women, respectively. The observed causal effects for hormones were maintained in the ER+ samples, but no effect was observed in the ER-samples. Some differences were also observed for reproductive traits, for example, age at first birth had a direct effect only on the ER-cases (younger women). Additionally, there was a null effect of age at menopause and increased number of births among ER-breast cancer cases, which is reasonable since those exposures are less prevalent in younger women who typically present with ER-breast tumours.

Although we used the largest GWAS available for each mediator trait of interest, many of these data sources possess specific limitations that may have prevented us from identifying the mediated effect. When investigating hormones as outcomes, we used data from UK Biobank, which is the same sample as our main exposure (childhood body size). To assess whether sample overlap could have potentially led to “winner’s curse” bias in step 1 of MR, a split-sample analysis was performed. Results of non-overlapping samples analysis were similar to the full sample analysis, suggesting that using the same data source for both exposure and outcome had little impact on our findings. Another limitation related to the hormones measures is that these were quantified in a predominantly post-menopausal sample of women (average age in UK Biobank is 56 years), where sex hormone levels are considerably different to those before menopause [61].

For the reproductive traits, we prioritised non-UK Biobank datasets in the main analysis to minimise the problem of sample overlap. While these datasets typically were from smaller sample sizes (and therefore fewer instruments), the directions of observed effects were consistent with the analyses performed on UK Biobank data (Supplementary Table S2).

The most inconclusive results were observed for glycaemic traits, likely due to smaller samples sizes and mixed-sex samples within these data sources. For traits with multiple available data sources, we prioritised those containing female-only participants [62], which reduced the power of the analyses but showed more relevant effects than in the mixed-sex analyses (Supplementary Tables S2 and S3).

The unavailability of full summary data for the physical traits of interest is a major constraint in this study. Since only the top GWAS hits from mammographic density studies by Sieh *et al* (2020) [48], Brand *et al* (2018) [50] and Lindström *et al* (2014) [49] were available, we were unable to estimate the effect of childhood body size on these traits and the extent to which they could mediate the relationship with breast cancer. High MD is a major breast cancer risk factor and, importantly, is therapeutically modifiable [63]. Moreover, higher adiposity in childhood and adolescence has been associated with lower MD throughout adulthood [24]. In light of several recent studies [64], [65] suggesting a plausible role of MD in the mediation of the protective effect of childhood body size on breast cancer risk, applying our MR framework to these datasets is an important aim for the future. We were also unable to perform the full analysis on breast size data. However, a previous study using MVMR showed that breast size is an unlikely to be a mediator of BMI effect on breast cancer risk Nick Sern Ooi *et al* (2019) [15].

While most of our analysis focused on mediators measured in adulthood, assessing mediators measured earlier in life would be useful for exploring the life-course effects of childhood body size. Investigating how childhood body size score influences plausible mediators over time, including an assessment of its effects on other anthropometric measures such as growth, changes in body composition and fat distribution [66], would provide another critical step to improve mechanistic understanding of its protective effect on breast cancer risk.

Another important point to raise is the gene-environment equivalence assumption in MR, i.e. that if childhood body size is influenced genetically or environmentally this will have the same effect on the outcome [67]. It is necessary to consider whether childhood adiposity produced by environmental/lifestyle factors can reduce the risk of breast cancer in the same way as has been estimated using genetic variants that affect body size in early life.

It is also important to mention that current MR methods for meditation analysis assume linear associations. However, it is possible that the effects of childhood body size and mediators are non-linear, which could lead to an apparent lack of mediation despite the presence of the true meditating effect. Additionally, two-step MR and MVMR assume no interaction between the exposure and the mediator on the outcome. When assumptions of linearity and no interaction are not satisfied, the magnitude of the estimated effect may be affected [68].

In summary, here we systematically reviewed a set of potential mediators for the observed protective effect of increased childhood body size on breast cancer risk. Individually, none of the tested traits were found to strongly mediate this effect. However, it is plausible that several related traits may collectively contribute to the mediated effect, which could be explored in multi-mediator MVMR analyses in future studies. It would also be interesting to explore mediation effects on breast cancer experienced pre- and post-menopause, and ultimately by molecular subtypes of the disease. Mediation may also occur via a pathway that we have not considered in our study, or via mammographic density, which could not be fully explored in this study due to the lack of available data. Finally, future work could explore proximal molecular mediators (e.g. breast tissue gene expression and methylation) to determine if early-life and adult adiposity have different effects on breast biology, which would be a critical step in deciphering the protective effect investigated in this work.

Our systematic investigation of mediators was designed as a prioritisation workflow, in which the initial MR analysis results (two-step MR) were used to select mediators for more advanced analyses (MVMR, mediation) based on the presence of causal effects and adequate performance in sensitivity analyses. Although we failed to identify a sole plausible mediator, we systematically report the MR results for the majority of obvious candidates from the largest available GWAS datasets to our knowledge. As new GWAS data becomes available, a similar approach can be applied, or used for investigating other biological questions (e.g. as shown in a recent study [69], aiming to identify the mediators of height effect on coronary artery disease). While we adopted a hypothesis-driven approach to investigate potential mediators, in future work, data mining platforms such as EpiGraphDB (epigraphdb.org) [70] may be used to facilitate the identification of novel mediator traits/biomarkers, or candidates for multi-mediator MVMR analyses.

## Methods

### Data Sources

UK Biobank [71] was the sources for the main exposure GWAS (childhood body size) and the Breast Cancer Association Consortium (BCAC) [51] was the main source for outcome GWAS (breast cancer) used in this work. The sources of the mediator trait GWAS are summarised in Table 1.

UK Biobank is a population-based health research resource consisting of approximately 500,000 people, aged between 38 years and 73 years, who were recruited between the years 2006 and 2010 from across the UK. A full description of the study design, participants and quality control (QC) methods have been described in detail previously [71]. The GWAS of childhood body size and adult body size used in this study were performed by Richardson *et al* (2020) [16] on UK Biobank data (N=246,511; female only).

BCAC breast cancer GWAS includes 228,951 samples (122,977 cases and 105,974 controls) of European ancestry. The cases include both oestrogen receptor-positive (N=69,501) and oestrogen receptor-negative (N=21,468) participants [51]. The details of cohort design and genotyping protocol are described elsewhere (bcac.ccge.medschl.cam.ac.uk, ccge.medschl.cam.ac.uk/research/consortia/icogs/). The BCAC data was accessed through OpenGWAS [36] (gwas.mrcieu.ac.uk) under the following IDs: *‘ieu-a-1126’* (full sample), *‘ieu-a-1127’* (ER+), *‘ieu-a-1128’* (ER-).

### Description of selected traits

Childhood body size is a categorical trait describing body size at age 10, with three categories (‘thinner than average’, ‘about average’, ‘plumper than average’), from a questionnaire completed by adult participants of UK Biobank. Adult body size measure was converted from continuous adult BMI in UK biobank into three groups based on the proportions of childhood body size data to ensure that the GWAS results of both measures are comparable [16]. The genetic scores for childhood and adult body size were independently validated in two separate cohorts (HUNT Study (Norway) [72] and Young Finns Study [73]), which confirmed that the genetic instruments extracted by Richardson *et al* (2020) [16] can reliably separate childhood and adult body size as distinct exposures.

Four groups of mediators were assessed: sex hormones, female reproductive traits, glycaemic traits, and physical traits.

1. Hormones: IGF-1 (Insulin-like growth factor 1), SHBG (sex hormone-binding globulin), oestradiol, testosterone (free, bioavailable, total). The different measures of testosterone were estimated as described in [55].
2. Reproductive traits: age at first birth, age at menarche (x2), number of births, age at menopause (x2).
3. Glycaemic traits: fasting insulin (x3), fasting glucose (x3), HbA1c (glycated haemoglobin A1c) (x2), HOMA-B (Homeostatic Model Assessment of β-cell function). HOMA-IR (insulin resistance) was considered in the analysis too, however, no robust instruments were identified.
4. Physical traits: breast size (x3), mammographic density (MD) (percent, dense/non-dense area) (x3). Full summary data was not available for breast size and MD, so only a small number of top GWAS hits were used in the analysis.

Several traits were available from multiple data sources (marked with xN). In the early stages of the analysis, all of them were evaluated, but only one version is presented in the final set of results. To prioritise a particular dataset over the rest we used the following criteria: 1) female-only sample, 2) non-UK Biobank data, 3) sample size. Table 1 shows the full set of tested datasets, highlighting the final selection with an asterisk.

### IEU GWAS pipeline

The GWAS of hormone mediators from UK Biobank were performed using the MRC IEU GWAS pipeline [35] which is based on BOLT-LMM (v2.3) [74] linear mixed model and an additive genetic model adjusted for sex, genotyping array, and 10 genetic principal components. The data were inverse rank normalised prior to the analysis; the results are quantified as standard deviation change.

### Mendelian randomization

Mendelian randomization (MR) is a type of instrumental variable (IV) analysis where genetic variants are used as proxies to uncover the causal relationship between a modifiable health exposure and a disease outcome [29]. There are three core assumptions that genetic variants need to satisfy to qualify as valid instruments for the causal inference: (1) variants have to be reliably associated with exposure of interest, (2) there cannot be any association with confounders affecting the exposure-outcome relationship, and (3) variants cannot be independently associated with the outcome, via pathway other than through the exposure (i.e. horizontal pleiotropy) [75].

The analyses in this work were performed using the two-sample (univariable) MR approach (Figure 3A), which relies on using GWAS summary statistics of two non-overlapping samples for exposure (sample 1) and outcome (sample 2) [76]. Two-sample MR is the basis for the more advanced analysis set-up described in the next sections.

Two-sample MR analyses were performed using the inverse-variance weighted (IVW) method [77], which is presented in the Results. Alongside IVW, other complementary MR methods were applied to assess the robustness of the causal estimates and to overcome any potential violations of the MR assumptions (e.g. horizontal pleiotropy) (see **Sensitivity analysis** for further details).

### Two-step MR

Two-step MR (also known as network MR) is a sequence of two (or more) MR analyses connected by a shared variable. Two-step analysis setup is used to assess whether an intermediate trait acts as a causal mediator between the main exposure and the outcome of interest [32], [78]. As shown in Figure 3B, in step 1, genetics variants (IVs) for the exposure are used to estimate the causal effect of the exposure variable on the potential mediator, then, in step 2, the mediator IVs are used to assess the causal effect of the mediator on the outcome [78]. The evidence of a causal effect in both steps suggests that the association between exposure and outcome is mediated by the intermediate variable to some extent (further details in **Mediation analysis**).

### MVMR

Multivariable Mendelian randomization (MVMR) is an extension to the standard univariable MR, which allows genetic variants to be associated with more than one exposure, and can estimate the direct causal effects of each exposure in a single analysis [33]. In this way, an exposure trait and a potential mediator can be analysed together to quantify the direct effect of both independently on the outcome (Figure 3C). The genetic variants included in the analysis have to be reliably associated with one or both exposures but not completely overlap (i.e. no perfect collinearity), and have to satisfy the MVMR-extended second and third assumptions of the standard MR analysis [34] [79]. Diagnostic methods and sensitivity tests for the robustness of MVMR estimates [79] [80] are described under **Sensitivity analysis**.

### MR analysis tools

All analyses were conducted using R (version 4.0.0). Univariate MR analyses and sensitivity tests were performed using the *TwoSampleMR* R package (version 0.5.4) [81], which was also used for accessing GWAS summary data deposited in OpenGWAS [36] (gwas.mrcieu.ac.uk). Multivariable MR was carried out by adapting *TwoSampleMR*’s functionality to be used on mixed data sources (see **Code availability**). Sensitivity analyses for multivariable MR were performed using *MVMR* R package (version 0.2) [34].

For all exposure and mediator datasets, the instruments were extracted from the full summary data by selecting SNPs under P-value < 5×10^−8^ threshed and clumping the resulting set of variants with r2 < 0.001. When extracting instruments from the outcome (breast cancer) GWAS summary statistics, unavailable SNPs were substituted by proxies with a minimum linkage disequilibrium r2 = 0.8. The rest of the settings were kept to defaults as per package version number.

### Sensitivity analysis

To further investigate the causal estimates found in the standard (IVW) MR analyses and to evaluate the validity of the analysed genetic instruments, MR-Egger [52] and weighted median MR [53] methods were used to overcome and accommodate for potential violations of the core MR assumptions. These complementary methods help to support the causal effects found with IVW, as a single method cannot account for all biological and statistical properties that may impact MR estimates. A variety of specialised sensitivity tests were applied, as recommended in [81].

To assess overall horizontal pleiotropy (violation of assumption 3), the intercept in the MR-Egger regression [52] was evaluated, and the heterogeneity among the genetic variants was quantified using the Cochran’s Q-statistic [82]. The intercept term in MR-Egger regression is a useful indication of whether directional horizontal pleiotropy is driving the results of an MR analysis. When the Egger intercept is close to zero (e.g. < 0.002) and the P-value is large, this can be interpreted as no evidence of a substantial directional (horizontal) pleiotropic effect. When the Q-statistic for heterogeneity (difference in individual ratio estimates) is high and the corresponding p-value is small, this suggests evidence for heterogeneity and possibly horizontal pleiotropy. However, this heterogeneity does not translate into a directional pleiotropic effect if the Egger intercept is (close to) zero (under the InSIDE assumption). A high Q-statistic can be also used as an indicator of one or more variant outliers in the analysis, which may also be violating the MR assumptions.

Additionally, scatter plots of SNP effects from exposure and outcome fitted by all tested MR methods were evaluated for any deviations which would also be indicative of heterogeneity and violations of MR assumptions. Single SNP forest plots were used summarise the effect of the exposure on the outcome due to each SNP separately, which is a helpful approach for visualising SNPs heterogeneity. Next, funnel plots were used to visually evaluate the direction of pleiotropy, which, if present, would be characterised by asymmetry in the plot. Finally, the sensitivity of causal inference to any individual genetic variant was tested by leave-one-out analysis, which is used to identify outliers.

In MVMR analyses, conditional F-statistics were used to evaluate the instrument strength [79], with F > 10 indicating suitable strength for the analysis. However, as in univariable MR, heterogeneity may be indicative of horizontal pleiotropy that does not act through one of the exposures. In MVMR, heterogeneity is quantified by Q_A_-statistic (also a further modification of Cochran’s Q), and small Q_A_ indicates a lack of directional pleiotropic effect [79].

To calculate both statistic measures, a phenotypic correlation matrix for each MVMR test had to be constructed. This was done by applying the method in for phenotypic matrix construction from GWAS summary data, available from *metaCCA* R package [83]. This is an alternative approach to using individual-level data for matrix construction. This was only applied in cases when both exposures in MVMR were from the same sample (i.e. UK Biobank). When data samples were different, the default settings for F and Q_A_ statistics were used (i.e. *gencov* parameter set to zero) [79].

In the cases where MVMR sensitivity tests indicated the presence of weak instruments and potential pleiotropy via heterogeneity, Q-minimisation approach (Q-het) for estimating causal association was used to supplement the estimated of MVMR-IVW approach [79].

### Split-sample analysis

One of the important requirements in two-sample MR is that the exposure and outcome GWAS are two non-overlapping datasets, which provides an advantage over the limitations in one-sample MR analysis of winner’s curse and anti-conservative weak instrument bias [84] [85]. In the two-sample MR analyses of childhood body size on hormones and some of the reproductive traits, this requirement was not satisfied, since the mediator GWAS were performed in the same cohort as the main exposure (childhood body size), i.e. UK Biobank.

To overcome the bias and evaluate the extent to which the results are affected, the MR analyses were repeated using a split-sample approach. This was possible due to the large sample size of the UK Biobank: the data were randomly split into two parts, and one part was used to carry out a new GWAS for the exposure (childhood body size), and the other part for the outcome (hormones/reproductive traits). With newly extracted instruments, step 1 of two-step MR and MVMR were repeated, with the resulting estimates are available in Supplementary Tables S15 and S16 and Supplementary Materials Section D. Finally, the requirement of having two separate samples does not apply to exposures used in MVMR, i.e. it is acceptable to use UK Biobank traits for both exposure and mediator used in the analysis [34].

### Mediation analysis

Mediation analysis is a method for quantifying the effects of an exposure on an outcome, which act directly, or indirectly via an intermediate variable (i.e. mediator) [68]. This analysis can identify which factors mediate the relationship between the exposure and the outcome enabling intervention on those mediators to mitigate or strengthen the effects of the exposure [86]. The “total” effect of exposure on outcome includes both “direct” effect and any “indirect” effects via one or more mediators.

In terms of MR, the total effect is captured by a standard univariable MR analysis (Figure 3A). To decompose direct and indirect effects, we use the results from two-step MR and MVMR (Figure 3B and 3C) in two mediation analysis methods: Difference method and Product method (Supplementary material Section E, Figure S5). For mediation analysis, it is important to have strong evidence of the total effect (Supplementary Table S1), effect in two-step MR and MVMR, and strong instruments in MVMR (measured by F-statistics) with no evidence of pleiotropy.

Lastly, although it is sometimes advised against calculating an indirect effect if the outcome is a binary variable (i.e. disease status) due to non-collapsibility of odds ratios [60], it has been shown that if the outcome effects have been quantified as log-odds ratios, it is acceptable to use betas of such outcome in the mediation analysis [68].

The main mediation method (Product method) and the indirect effect SE/CI estimation approach (Delta method) were chosen with the help of simulation analysis described in Supplementary Materials Section E.

## Data Availability

The availability of all data analysed in this study has been referenced throughout the manuscript and supplementary materials.

## Code and data availability

MR analysis code is available at https://github.com/mvab/mendelian-randomization-breast-cancer

Simulation analysis for selecting mediation method is available at https://github.com/mvab/simulation_for_MR_mediation

Hormones GWAS data generated with the IEU GWAS pipeline will be uploaded to OpenGWAS.

## Acknowledgements

This research has been conducted using the UK Biobank Resource under application number 15825.

## Funding

BLL – University of Bristol (Vice-Chancellor’s fellowship) and the Academy of Medical Sciences.

RCR – de Pass Vice Chancellor’s research fellowship at the University of Bristol

MV – University of Bristol Alumni Fund: Professor Sir Eric Thomas Scholarship

MV, GDS, ES, TGR, RR work in the Medical Research Council Integrative Epidemiology Unit at the University of Bristol supported by Medical Research Council (MC_UU_00011/1, MC_UU_00011/2 and MC_UU_00011/4).

This work was also supported by a Cancer Research UK programme grant (the Integrative Cancer Epidemiology Programme) (C18281/A19169).

## Competing interests

TGR is employed part-time by Novo Nordisk outside of this work. All other authors declare no competing interests.

## Notes

### Author Declarations

All analyses conducted in this study are based on data from summary-level datasets (whose ethical approval can be found in the corresponding studies referenced) or the UK Biobank study (Research Ethics Committee approval number: 11/NW/0382, app #15825).

## References

[1] H. Sung et al., “Global cancer statistics 2020: GLOBOCAN estimates of incidence and mortality worldwide for 36 cancers in 185 countries,” CA. Cancer J. Clin., vol. 0, no. 0, pp. 1–41, 2021.

[2] IARC, “International Agency for Research on Cancer: Estimated cumulative risk of incidence in 2020, in females, in high-income countries, by cancer site; based on GLOBOSCAN 2020 data,” 2021. [Online]. Available: https://gco.iarc.fr/today/online-analysis-multi-bars?v=2020&mode=cancer&mode_population=countries&population=900&populations=986&key=cum_risk&sex=2&cancer=39&type=0&statistic=5&prevalence=0&population_group=0&ages_group%5B%5D=0&ages_group%5B%5D=14&nb_item. [Accessed: 12-May-2021].

[3] S. A. Narod, J. Iqbal, and A. B. Miller, “Why have breast cancer mortality rates declined?,” J. Cancer Policy, vol. 5, pp. 8–17, Sep. 2015.

[4] N. Harbeck et al., Breast cancer, vol. 5, no. 1. 2019.

[5] K. L. Britt, J. Cuzick, and K. A. Phillips, “Key steps for effective breast cancer prevention,” Nat. Rev. Cancer, 2020.

[6] B. Lauby-Secretan, C. Scoccianti, D. Loomis, Y. Grosse, F. Bianchini, and K. Straif, “Body Fatness and Cancer — Viewpoint of the IARC Working Group,” N. Engl. J. Med., vol. 375, no. 8, pp. 794–798, Aug. 2016.

[7] K. Bhaskaran, I. Douglas, H. Forbes, I. Dos-Santos-Silva, D. A. Leon, and L. Smeeth, “Body-mass index and risk of 22 specific cancers: A population-based cohort study of 5·24 million UK adults,” Lancet, vol. 384, no. 9945, pp. 755–765, Aug. 2014.

[8] K. Liu et al., “Association between body mass index and breast cancer risk: Evidence based on a dose– response meta-analysis,” Cancer Manag. Res., vol. 10, pp. 143–151, 2018.

[9] C. M. Friedenreich, “Review of anthropometric factors and breast cancer risk,” Eur. J. Cancer Prev., vol. 10, no. 1, pp. 15–32, 2001.

[10] A. G. Renehan, M. Zwahlen, and M. Egger, “Adiposity and cancer risk: New mechanistic insights from epidemiology,” Nature Reviews Cancer, vol. 15, no. 8. Nature Publishing Group, pp. 484–498, 27-Aug-2015.

[11] H. J. Baer, S. S. Tworoger, S. E. Hankinson, and W. C. Willett, “Body Fatness at Young Ages and Risk of Breast Cancer Throughout Life,” vol. 171, no. 11, 2010.

[12] A. Furer et al., “Adolescent obesity and midlife cancer risk: a population-based cohort study of 2·3 million adolescents in Israel,” Lancet Diabetes Endocrinol., vol. 8, no. 3, pp. 216–225, 2020.

[13] A. G. Renehan et al., “Young adulthood body mass index, adult weight gain and breast cancer risk: the PROCAS Study (United Kingdom),” Br. J. Cancer, vol. 122, no. 10, pp. 1552–1561, 2020.

[14] Y. Guo et al., “Genetically Predicted Body Mass Index and Breast Cancer Risk: Mendelian Randomization Analyses of Data from 145,000 Women of European Descent,” PLoS Med., vol. 13, no. 8, p. e1002105, Aug. 2016.

[15] B. Nick Sern Ooi et al., “The genetic interplay between body mass index, breast size and breast cancer risk: a Mendelian randomization analysis,” Int. J. Epidemiol., pp. 781–794, 2019.

[16] T. G. Richardson, E. Sanderson, B. Elsworth, K. Tilling, and G. D. Smith, “Use of genetic variation to separate the effects of early and later life adiposity on disease risk: Mendelian randomisation study,” BMJ, vol. 369, 2020.

[17] E. M. Poole, S. S. Tworoger, S. E. Hankinson, E. S. Schernhammer, M. N. Pollak, and H. J. Baer, “Body size in early life and adult levels of insulin-like growth factor 1 and insulin-like growth factor binding protein 3,” Am. J. Epidemiol., vol. 174, no. 6, pp. 642–651, 2011.

[18] M. J. Schoemaker et al., “Association of Body Mass Index and Age with Subsequent Breast Cancer Risk in Premenopausal Women,” JAMA Oncol., vol. 4, no. 11, pp. e181771–e181771, Nov. 2018.

[19] C. Group on Hormonal Factors in Breast Cancer, “Menarche, menopause, and breast cancer risk: individual participant meta-analysis, including 118 964 women with breast cancer from 117 epidemiological studies Collaborative Group on Hormonal Factors in Breast Cancer*,” Lancet Oncol., vol. 13, pp. 1141–1151, 2012.

[20] G. V. Dall and K. L. Britt, “Estrogen effects on the mammary gland in early and late life and breast cancer risk,” Frontiers in Oncology, vol. 7, no. MAY. Frontiers Media S.A., p. 1, 26-May-2017.

[21] C. S. Berkey, J. D. Gardner, A. Lindsay Frazier, and G. A. Colditz, “Relation of childhood diet and body size to menarche and adolescent growth in girls,” Am. J. Epidemiol., vol. 152, no. 5, pp. 446–452, 2000.

[22] L. Hilakivi-Clarke et al., “Tallness and overweight during childhood have opposing effects on breast cancer risk,” Br. J. Cancer, vol. 85, no. 11, pp. 1680–1684, Nov. 2001.

[23] A. Pettersson and R. M. Tamimi, “Breast fat and breast cancer,” Breast Cancer Research and Treatment, vol. 135, no. 1. Breast Cancer Res Treat, pp. 321–323, Aug-2012.

[24] L. Yochum, R. M. Tamimi, and S. E. Hankinson, “Birthweight, early life body size and adult mammographic density: a review of epidemiologic studies,” Cancer Causes and Control, vol. 25, no. 10. Kluwer Academic Publishers, pp. 1247–1259, 11-Oct-2014.

[25] A. G. Ghadge, P. Dasari, J. Stone, E. W. Thompson, R. L. Robker, and W. V. Ingman, “Pubertal mammary gland development is a key determinant of adult mammographic density,” Semin. Cell Dev. Biol., no. July, 2020.

[26] P. Boyle et al., “Blood glucose concentrations and breast cancer risk in women without diabetes: A meta-analysis,” Eur. J. Nutr., vol. 52, no. 5, pp. 1533–1540, Aug. 2013.

[27] A. V Hernandez, M. Guarnizo, Y. Miranda, V. Pasupuleti, and A. Deshpande, “Association between Insulin Resistance and Breast Carcinoma: A Systematic Review and Meta-Analysis,” PLoS One, vol. 9, no. 6, p. 99317, 2014.

[28] X. Shu et al., “Associations of obesity and circulating insulin and glucose with breast cancer risk: A Mendelian randomization analysis,” Int. J. Epidemiol., vol. 48, no. 3, pp. 795–806, 2019.

[29] S. Ebrahim and G. Davey Smith, “Mendelian randomization: Can genetic epidemiology help redress the failures of observational epidemiology?,” Int. J. Epidemiol., vol. 32, no. 1, pp. 1–22, Feb. 2003.

[30] G. Davey Smith and G. Hemani, “Mendelian randomization: Genetic anchors for causal inference in epidemiological studies,” Hum. Mol. Genet., vol. 23, no. R1, pp. R89–R98, Sep. 2014.

[31] D. A. Lawlor, “Commentary: Two-sample Mendelian randomization: Opportunities and challenges,” Int. J. Epidemiol., vol. 45, no. 3, pp. 908–915, 2016.

[32] C. L. Relton and G. Davey Smith, “Two-step epigenetic mendelian randomization: A strategy for establishing the causal role of epigenetic processes in pathways to disease,” Int. J. Epidemiol., vol. 41, no. 1, pp. 161–176, 2012.

[33] S. Burgess and S. G. Thompson, “Multivariable Mendelian randomization: The use of pleiotropic genetic variants to estimate causal effects,” Am. J. Epidemiol., vol. 181, no. 4, pp. 251–260, 2015.

[34] E. Sanderson, G. Davey Smith, F. Windmeijer, and J. Bowden, “An examination of multivariable Mendelian randomization in the single-sample and two-sample summary data settings,” Int. J. Epidemiol., vol. 48, no. 3, pp. 713–727, Jun. 2019.

[35] B. Elsworth, R. Mitchell, C. Raistrick, L. Paternoster, G. Hemani, and T. Gaunt, “MRC IEU UK Biobank GWAS pipeline version 2. University of Bristol.” 2019.

[36] B. Elsworth et al., “The MRC IEU OpenGWAS data infrastructure,” bioRxiv, p. 2020.08.10.244293, Aug. 2020.

[37] A. Buniello et al., “The NHGRI-EBI GWAS Catalog of published genome-wide association studies, targeted arrays and summary statistics 2019,” Nucleic Acids Res., vol. 47, no. D1, pp. D1005–D1012, Jan. 2019.

[38] J. R. B. Perry et al., “Parent-of-origin-specific allelic associations among 106 genomic loci for age at menarche,” Nature, vol. 514, no. 7520, pp. 92–97, Oct. 2014.

[39] F. R. Day et al., “Large-scale genomic analyses link reproductive aging to hypothalamic signaling, breast cancer susceptibility and BRCA1-mediated DNA repair,” Nat. Genet., vol. 47, no. 11, pp. 1294–1303, Nov. 2015.

[40] V. Lagou et al., “Sex-dimorphic genetic effects and novel loci for fasting glucose and insulin variability,” Nat. Commun., vol. 12, no. 1, pp. 1–18, Dec. 2021.

[41] A. K. Manning et al., “A genome-wide approach accounting for body mass index identifies genetic variants influencing fasting glycemic traits and insulin resistance,” Nat. Genet., vol. 44, no. 6, pp. 659–669, Jun. 2012.

[42] R. A. Scott et al., “Large-scale association analyses identify new loci influencing glycemic traits and provide insight into the underlying biological pathways,” Nat. Genet., vol. 44, no. 9, pp. 991–1005, Sep. 2012.

[43] E. Wheeler et al., “Impact of common genetic determinants of Hemoglobin A1c on type 2 diabetes risk and diagnosis in ancestrally diverse populations: A transethnic genome-wide meta-analysis,” PLoS Med., vol. 14, no. 9, Sep. 2017.

[44] N. Soranzo et al., “Common variants at 10 genomic loci influence hemoglobin A1C levels via glycemic and nonglycemic pathways,” Diabetes, vol. 59, no. 12, pp. 3229–3239, Dec. 2010.

[45] J. Dupuis et al., “New genetic loci implicated in fasting glucose homeostasis and their impact on type 2 diabetes risk,” Nat. Genet., vol. 42, no. 2, pp. 105–116, 2010.

[46] J. K. Pickrell, T. Berisa, J. Z. Liu, L. Ségurel, J. Y. Tung, and D. A. Hinds, “Detection and interpretation of shared genetic influences on 42 human traits,” Nat. Genet., vol. 48, no. 7, pp. 709–717, Jul. 2016.

[47] N. Eriksson et al., “Genetic variants associated with breast size also influence breast cancer risk,” BMC Med. Genet., vol. 13, Jun. 2012.

[48] W. Sieh et al., “Identification of 31 loci for mammographic density phenotypes and their associations with breast cancer risk,” Nat. Commun., vol. 11, no. 1, Dec. 2020.

[49] S. Lindström et al., “Genome-wide association study identifies multiple loci associated with both mammographic density and breast cancer risk,” Nat. Commun., vol. 5, p. 5303, Oct. 2014.

[50] J. S. Brand, K. Humphreys, J. Li, R. Karlsson, P. Hall, and K. Czene, “Common genetic variation and novel loci associated with volumetric mammographic density,” Breast Cancer Res., vol. 20, no. 1, Apr. 2018.

[51] K. Michailidou et al., “Association analysis identifies 65 new breast cancer risk loci,” Nature, vol. 551, no. 7678, pp. 92–94, Nov. 2017.

[52] J. Bowden, G. D. Smith, and S. Burgess, “Mendelian randomization with invalid instruments: Effect estimation and bias detection through Egger regression,” Int. J. Epidemiol., vol. 44, no. 2, pp. 512–525, May 2015.

[53] J. Bowden, G. Davey Smith, P. C. Haycock, and S. Burgess, “Consistent Estimation in Mendelian Randomization with Some Invalid Instruments Using a Weighted Median Estimator,” Genet. Epidemiol., vol. 40, no. 4, pp. 304–314, May 2016.

[54] N. Murphy et al., “Insulin-like growth factor-1, insulin-like growth factor-binding protein-3, and breast cancer risk: observational and Mendelian randomization analyses with ∼ 430 000 women,” Ann. Oncol., vol. 31, no. 5, pp. 641–649, 2020.

[55] K. S. Ruth et al., “Using human genetics to understand the disease impacts of testosterone in men and women,” Nat. Med., vol. 26, no. 2, pp. 252–258, Feb. 2020.

[56] N. L. Dimou et al., “Sex hormone binding globulin and risk of breast cancer: A Mendelian randomization study,” Int. J. Epidemiol., vol. 48, no. 3, pp. 807–816, 2019.

[57] T. J. Key et al., “Insulin-like growth factor 1 (IGF1), IGF binding protein 3 (IGFBP3), and breast cancer risk: Pooled individual data analysis of 17 prospective studies,” Lancet Oncol., vol. 11, no. 6, pp. 530–542, Jun. 2010.

[58] P. F. Christopoulos, P. Msaouel, and M. Koutsilieris, “The role of the insulin-like growth factor-1 system in breast cancer,” Molecular Cancer, vol. 14, no. 1. BioMed Central Ltd., p. 43, 15-Feb-2015.

[59] H. Yu and T. Rohan, “Role of the insulin-like growth factor family in cancer development and progression,” Journal of the National Cancer Institute, vol. 92, no. 18. pp. 1472–1489, 20-Sep-2000.

[60] S. Burgess, D. J. Thompson, J. M. B. Rees, F. R. Day, J. R. Perry, and K. K. Ong, “Dissecting causal pathways using mendelian randomization with summarized genetic data: Application to age at menarche and risk of breast cancer,” Genetics, vol. 207, no. 2, pp. 481–487, Oct. 2017.

[61] E. R. Simpson et al., “Estrogen—the Good, the Bad, and the Unexpected,” Endocr. Rev., vol. 26, no. 3, pp. 322–330, May 2005.

[62] V. Lagou, R. Mägi, and J.-J. J. Hottenga, “Fasting glucose and insulin variability: sex-dimorphic genetic effects and novel loci,” Prep., 2019.

[63] J. Cuzick et al., “Tamoxifen-induced reduction in mammographic density and breast cancer risk reduction: A nested case-control study,” J. Natl. Cancer Inst., vol. 103, no. 9, pp. 744–752, May 2011.

[64] J. L. Hopper et al., “Childhood body mass index and adult mammographic density measures that predict breast cancer risk,” Breast Cancer Res. Treat., vol. 156, no. 1, pp. 163–170, 2016.

[65] Y. Han et al., “Adiposity Change Over the Life Course and Mammographic Breast Density in Postmenopausal Women,” Cancer Prev. Res., vol. 13, no. 5, pp. 475–482, 2020.

[66] R. J. F. Loos and T. O. Kilpeläinen, “Genes that make you fat, but keep you healthy,” Journal of Internal Medicine, vol. 284, no. 5. Blackwell Publishing Ltd, pp. 450–463, 01-Nov-2018.

[67] G. Davey Smith, “Epigenesis for epidemiologists: Does evo-devo have implications for population health research and practice?,” Int. J. Epidemiol., vol. 41, no. 1, pp. 236–247, Feb. 2012.

[68] A. R. Carter et al., “Mendelian randomisation for mediation analysis: Current methods and challenges for implementation,” European Journal of Epidemiology. 2021.

[69] E. Marouli et al., “Mendelian randomisation analyses find pulmonary factors mediate the effect of height on coronary artery disease,” Commun. Biol., vol. 2, no. 1, pp. 1–9, Dec. 2019.

[70] Y. Liu et al., “EpiGraphDB: a database and data mining platform for health data science,” Bioinformatics, Nov. 2020.

[71] C. Sudlow et al., “UK Biobank: An Open Access Resource for Identifying the Causes of a Wide Range of Complex Diseases of Middle and Old Age,” PLoS Med., vol. 12, no. 3, Mar. 2015.

[72] M. Brandkvist et al., “Separating the genetics of childhood and adult obesity: a validation study of genetic scores for body mass index in adolescence and adulthood in the HUNT Study,” Hum. Mol. Genet., vol. 29, no. 24, pp. 3966–3973, Dec. 2020.

[73] T. G. Richardson et al., “Evaluating the direct effects of childhood adiposity on adult systemic metabolism: a multivariable Mendelian randomization analysis,” Int. J. Epidemiol., Mar. 2021.

[74] P. R. Loh et al., “Efficient Bayesian mixed-model analysis increases association power in large cohorts,” Nat. Genet., vol. 47, no. 3, pp. 284–290, Feb. 2015.

[75] D. Lawlor, “Mendelian randomization: Using genes as instruments for making causal inferences in epidemiology,” no. April, pp. 4267–4278, 2008.

[76] S. Burgess, R. A. Scott, N. J. Timpson, G. D. Smith, and S. G. Thompson, “Using published data in Mendelian randomization: A blueprint for efficient identification of causal risk factors,” Eur. J. Epidemiol., vol. 30, no. 7, pp. 543–552, Jul. 2015.

[77] S. Burgess, A. Butterworth, and S. G. Thompson, “Mendelian randomization analysis with multiple genetic variants using summarized data,” Genet. Epidemiol., vol. 37, no. 7, pp. 658–665, Nov. 2013.

[78] J. Zheng et al., “Recent Developments in Mendelian Randomization Studies,” Curr. Epidemiol. Reports, vol. 4, no. 4, pp. 330–345, 2017.

[79] E. Sanderson, W. Spiller, and J. Bowden, “Testing and Correcting for Weak and Pleiotropic Instruments in Two-Sample Multivariable Mendelian Randomisation,” bioRxiv, pp. 1–31, 2020.

[80] J. M. B. Rees, A. M. Wood, and S. Burgess, “Extending the MR-Egger method for multivariable Mendelian randomization to correct for both measured and unmeasured pleiotropy,” Stat. Med., vol. 36, no. 29, pp. 4705– 4718, Dec. 2017.

[81] G. Hemani et al., “The MR-base platform supports systematic causal inference across the human phenome,” Elife, vol. 7, May 2018.

[82] J. Bowden et al., “Improving the accuracy of two-sample summary-data Mendelian randomization: Moving beyond the NOME assumption,” Int. J. Epidemiol., vol. 48, no. 3, pp. 728–742, 2019.

[83] A. Cichonska, “metaCCA: Package for summary statistics-based multivariate meta-analysis of genome-wide association studies using canonical correlation analysis,” 2019.

[84] J. Yarmolinsky et al., “Causal inference in cancer epidemiology: What is the role of mendelian randomization?,” Cancer Epidemiology Biomarkers and Prevention, vol. 27, no. 9. American Association for Cancer Research Inc., pp. 995–1010, 01-Sep-2018.

[85] P. C. Haycock, S. Burgess, K. H. Wade, J. Bowden, C. Relton, and G. D. Smith, “Best (but oft-forgotten) practices: The design, analysis, and interpretation of Mendelian randomization studies,” American Journal of Clinical Nutrition, vol. 103, no. 4. American Society for Nutrition, pp. 965–978, 01-Apr-2016.

[86] E. Sanderson, “Multivariable Mendelian Randomization and Mediation,” Cold Spring Harb. Perspect. Med., p. a038984, 2020.

