## Supplementary Materials for "Deciphering how early life adiposity influences breast cancer risk using Mendelian randomization"

#### Tables file summary

**Supplementary_Tables_1-18.xls**

S1 *reproduction of Richardson et al 2020 paper results:* MR and MVMR results: effects of childhood body size and adult body size on breast cancer

S2 step 1 MR results (IVW, MR-Egger, Median), full BC sample
S3 step 2 MR results (IVW, MR-Egger, Median), full BC sample
S4 MVMR (childhood body size + mediator), full BC sample
S5 Mediator prioritisation decision table *(in this document)*

S6 step 1 sensitivity tests (Egger intercept, Cochran's Q), full BC sample
S7 step 2 sensitivity tests (Egger intercept, Cochran's Q), full BC sample
S8 MVMR (childhood body size, mediator) sensitivity tests (F-statistics, QA), full BC sample

S9 step 2 MR results (IVW, MR-Egger, Median), ER+ BC sample
S10 step 2 MR results (IVW, MR-Egger, Median), ER- BC sample
S11 MVMR results (childhood body size, mediator), ER+ BC sample
S12 MVMR results (childhood body size, mediator), ER- BC sample

S13 MVMR results (childhood body size, adult body size, mediator), full BC sample
S14 MVMR results (childhood body size, childhood height, mediator), full BC sample

S15 Split sample analysis; MR step1 (IVW)
S16 Split sample analysis; MVMR, full BC sample

S17 Mediation analysis results
S18 Simulation analysis results: sd of 1000 betas and mean of 1000 SE, for each beta/se method

#### Supplementary materials summary

Materials in **this** document:

Section A: Mediator prioritisation chart and table S5

Section B: Sensitivity analysis results

Section C: Split sample results

Section D: Additional MR analyses
- Total vs direct effect
- MVMR with extra traits

Section E: Mediation analysis details
- Mediation methods
- Standard error estimation
- Mediation analysis full results
- Simulation analysis for selecting the approach for mediation analysis
- Proportion mediated calculation

### Supplementary Materials

#### Section A. Mediator prioritisation

Mediator prioritisation can be visualised in two ways – decision tree (Figure S1) and decision table (Table S5). Both show the conditions/performance that a mediator analysis has to meet/show to be ‘qualified’ to be analyses in further steps that require the data that to be robust enough.

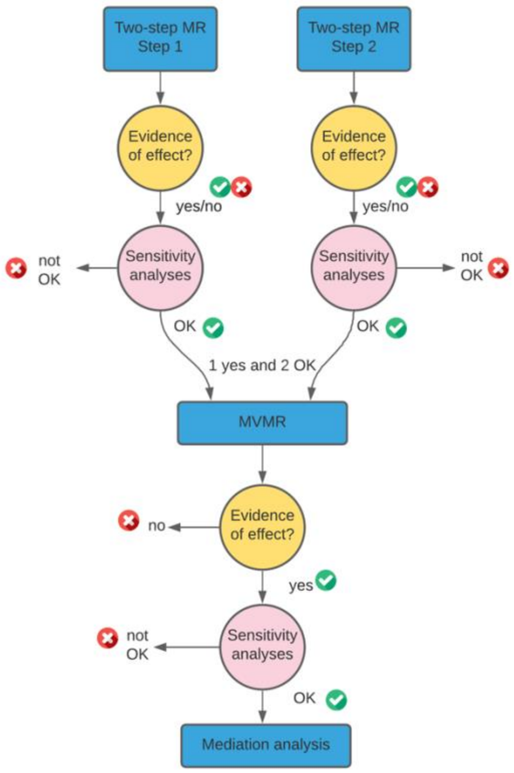


**Figure S1. Mediator prioritisation decision tree.** *For MVMR*: at least one step from two-step MR has to have evidence of effect, and both have to show adequate sensitivity tests results. *For mediation*: MVMR effect and satisfactory F-statistics (sensitivity analysis).

**Supplementary Table S5. Mediator prioritisation decision table. “**Effect” is evidence of a causal effect in the analysis (1 - present, 0 - not); “Sensitivity/F-stats” indicates consistent and satisfactory performance in the sensitivity analyses (1 – yes, 0 – no); “Do MVMR/mediation?” indicates whether the test was selected/prioritised to be done based on effect/sensitivity. *For MVMR*: at least one step has ‘effect’, and both show adequate sensitivity tests results. *For mediation*: MVMR effect and satisfactory F-statistics.

| | Step 1 effect | Step1 sensitivity | Step 2 effect | Step2 sensitivity | Do MVMR? | MVMR effect | F-stats | Do mediation? |
|---|---|---|---|---|---|---|---|---|
| IGF-1 | 1 | 1 | 1 | 1 | yes | 1 | 1 | yes |
| SHBG | 1 | 1 | 0 | 1 | yes | 0 | 1 | no |
| Oestradiol | 1 | 0 | 0 | 0 | no | | | no |
| Testosterone (bio) | 1 | 1 | 1 | 1 | yes | 1 | 1 | yes |
| Testosterone(free) | 1 | 1 | 1 | 1 | yes | 1 | 1 | yes |
| Testosterone(total) | 0 | 1 | 1 | 1 | yes | 1 | 1 | yes |
| Age at first birth | 1 | 1 | 0 | 0 | no | | | no |
| Age at menarche | 1 | 1 | 0 | 1 | yes | 1 | 1 | yes |
| Age at menopause | 0 | 1 | 1 | 1 | yes | 1 | 1 | yes |
| Number of births | 0 | 1 | 0 | 0 | no | | | no |
| Fasting glucose | 0 | 1 | 0 | 1 | no | 0 | 1 | no |
| Fasting insulin | 1 | 1 | 0 | 0 | no | | | no |
| Hb1ac | 1 | 1 | 0 | 1 | yes | 0 | 1 | no |
| HOMA-B | 1 | 1 | 0 | 0 | no | | | no |

#### Section B. Sensitivity analysis results

The sensitivity analyses of step 1 of two-step MR analyses (effect of childhood body size on mediators) showed consistent results across all mediator groups (Supplementary Table S6). The direction of estimates was the same for all tested mediators, and the magnitude of the estimate was comparable across three MR methods, except for a higher MR-Egger effect observed for SHBG. The MR-Egger intercepts were close to zero (< +/-0.003) with large p-values (> 0.2), indicating little evidence of directional pleiotropy, except for fasting insulin, which had intercept p-value of 0.006. Cochran's Q values were large with p-values (<0.0001) for hormones and reproductive traits, suggesting the presence of heterogeneity. Considerably smaller Q-values and large Q p-value were observed for all glycaemic traits and oestradiol, indicating little evidence for heterogeneity.

The sensitivity analyses of step 2 of two-step MR analyses (effect of mediators on breast cancer) showed less robust results for some mediator groups (Supplementary Table S7). Among the hormones, the direction of effect was consistent across all three MR methods, and the Egger intercept was small (close to zero) with a large p-value. Cochran's Q p-values were also small, indicating little evidence for heterogeneity in the hormones group. The exception was oestradiol, for which only one genome-wide significant SNP was available, meaning only a single Wald ratio test could be performed, and its effect has to be interpreted with caution. For reproductive traits, the direction of the effects was consistent, and the intercepts were sufficiently close to zero, but MR-Egger produced wider confidence intervals in all cases. None of the glycaemic traits showed evidence of the effect in IVW, and horizontal pleiotropy detected with other methods suggests likely bias of the estimates towards the null. For fasting insulin, MR-Egger analysis showed a strong effect in the opposite direction (observed on two out of three data sources, although based on < 5 SNPs), with Egger intercept for the main data source being away from zero (0.055). Similarly, HOMA-B had a limited number of instruments, and sensitivity tests highlighted its potentially biased and inconclusive results. Fasting glucose and Hb1ac performed adequately in the sensitivity tests. Finally, in the physical traits group, as only a limited number of instruments was available, as expected, most sensitivity analyses detected problems.

#### Section C. Additional MVMR analyses

##### Total vs direct effect of selected mediators on breast cancer

Figure S2 shows the total and direct effect of mediators on breast cancer. For all mediators (except age at menarche), the effects are very similar. In the case of age at menarche, in univariable MR, there was little evidence of total effect, while in MVMR, the direct effect (i.e. effect accounted for childhood body size) point estimate was notably more negative with CIs showing the evidence of this effect. The estimates are available in Tables S3-S4.

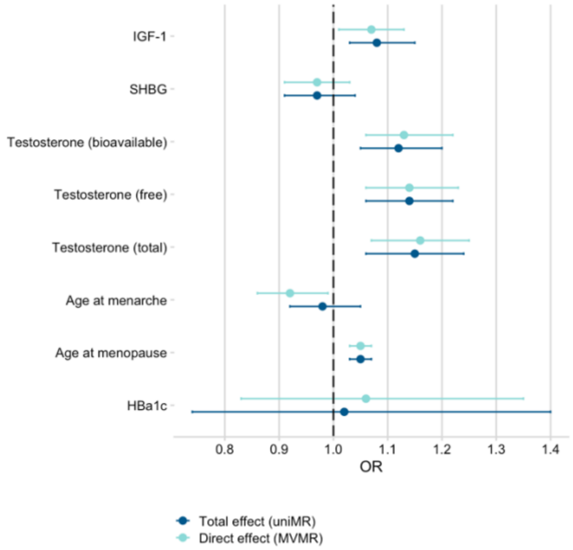


**Figure S2. Comparison of total and direct effect of a subset of mediators on breast cancer.** Total effect was estimated with univariable MR (step 2) and direct effect was estimated in MVMR with childhood body size effect taken into the account. Bars indicate 95% confidence intervals around the point estimates from IVW analyses.

##### MVMR with additional variables

The main MVMR analysis in this project aimed to separate the effects of childhood body size and a selected mediator. However, it was also important to consider if any effect can be explained by other related anthropometric measures. Therefore, we carried out additional MVMR analyses where we added adult BMI and childhood height as the additional variable in the MVMR model (two separate analyses). Figure S3 shows the direct effects of mediators on breast cancer from the three models.

When accounted for the effect of adult body size, the mediator effect estimates were not affected considerably compared to childhood body size. However, accounting for childhood height reduced the direct estimates for most mediators and attenuated IGF-1 and age at menarche effects to overlap the null. The estimates are available in Tables S5, S13, S14.

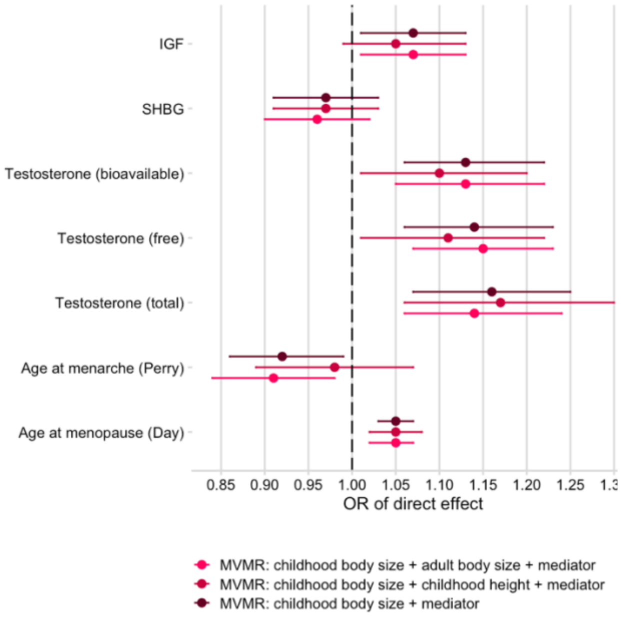


**Figure S3. Comparison of direct effect of mediators on breast cancer while accounting for childhood body size and other traits.** The MVMR models account for (1) childhood body size only (main MVMR analysis), (2) childhood body size and adult body size, and (3) childhood body size and childhood height.

#### Section D. Split-sample results

Figure S4 shows the MR results from full-sample and split-sample analyses of childhood body size (exposure) and selected hormones and reproductive traits (outcomes). For hormones, the effect size and CIs were very similar between the two analyses. In the split-sample analysis of reproductive traits, we observed larger effect sizes with wider CIs on age at menarche, age at first birth, and age at menarche. Overall, the sample overlap in the full-sample analysis does not seem to be upwardly biasing estimates, on the contrary, it seems to produce more conservative estimates. Therefore, we conclude that, in this study, using the same sample for both exposure and outcome has not introduced any substantial bias in the effect estimates.

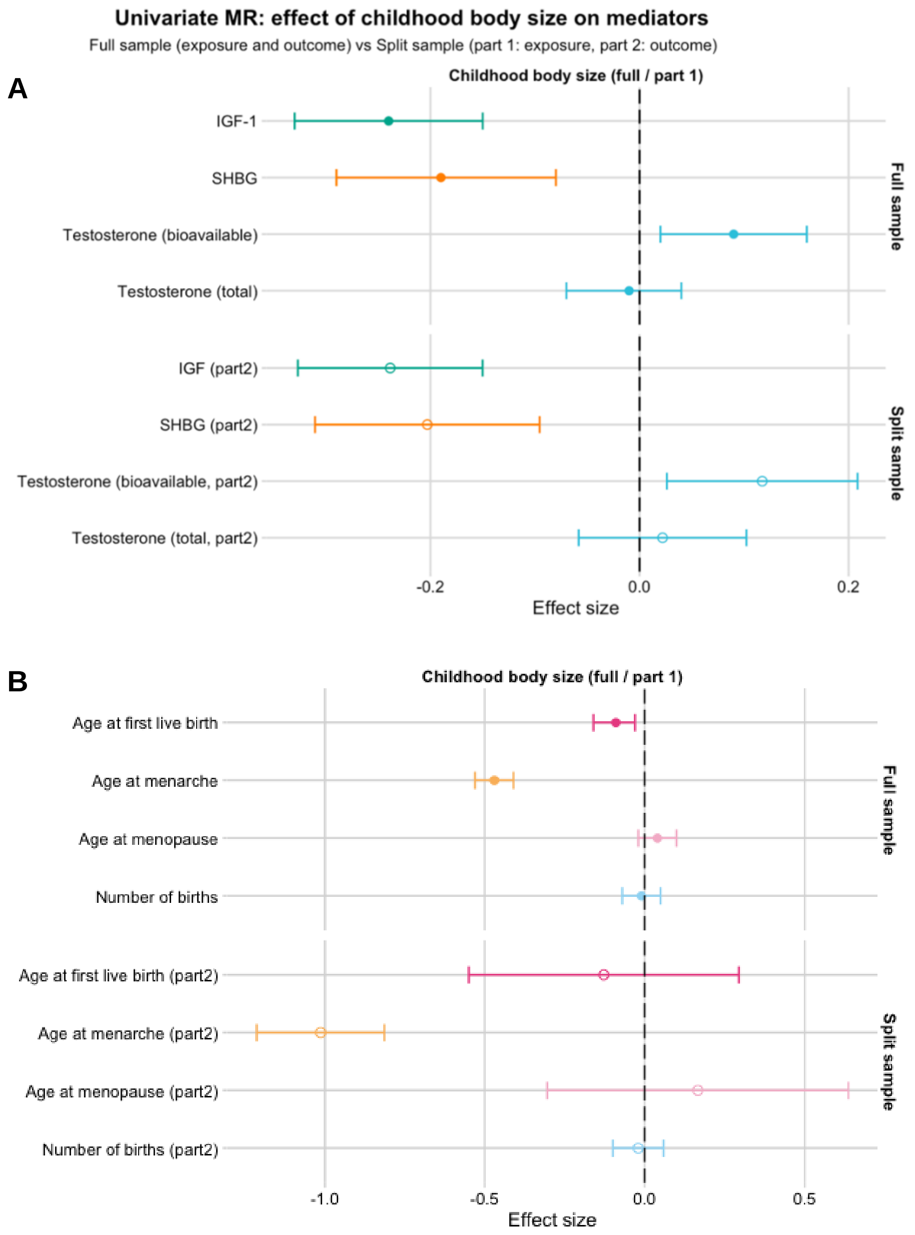


**Figure S4. Comparison of childhood body size effect estimates on hormones (A) and reproductive traits (B) from the full (top) and split sample (bottom) analyses.** In the full-sample analysis, the entire UK Biobank cohort was used to extract exposure instruments (childhood body size), and it was used as the other GWAS (hormones / reproductive traits). In the split-sample analysis, the cohort was randomly split into parts, and part 1 was used to extract exposure instruments, while part 2 was used as the outcome GWAS.

#### Section E. Mediation analysis details

##### Mediation methods

As outlined in Figure S5, in Difference method, to estimate the indirect effect, we subtract the direct effect of exposure on the outcome from MVMR (model with the mediator of interest) from the total effect of exposure on the outcome (univariable MR) [1]. In Product method (also known as 'product of coefficients'), we multiply the results from two steps of two-step MR analysis (i.e., the effect of exposure on the mediator and the effect of the mediator on the outcome) to get the indirect effect [2], [3]. It is sometimes recommended to replace the second term with the direct effect of the mediator on the outcome from MVMR analysis [4], but the results should be similar if there is sufficient instrument strength. In this paper, the direct effect was used in the analysis.

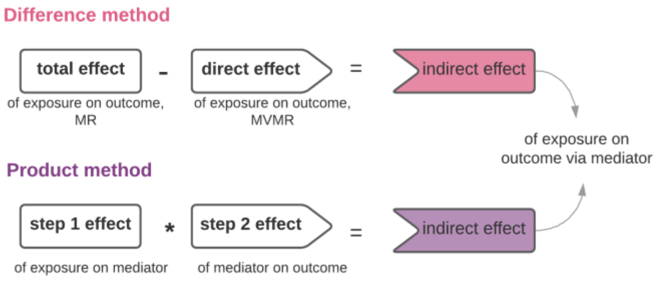


**Figure S5. Mediation analysis methods.** *Difference method:* the indirect effect is estimated as a difference between total and direct effect of exposure on the outcome, from univariable MR and multivariable MR, respectively. *Product method:* the indirect effect is estimated as the product of coefficients of the total effect of exposure on the mediator (step 1 of two-step MR) and total effect of mediator on the outcome (step 2 of two-step MR). The second term can be replaced by the direct effect of a mediator on the outcome from multivariable MR.

##### Standard error estimation methods

For the estimation of the confidence intervals (CIs) of the indirect effect, standard error (SE) has to be calculated. For each mediation method, there is a corresponding SE calculation method. To calculate SE of indirect effect from Difference method, 'Propagation of errors' (PoE) is used, as outlined in [1]. For Product method indirect effect SE calculation, we can use Delta method (also known as "Sobel test" [5]) or a simplified version of PoE.

*"Delta method"* – can be applied to product method results

```
se.indirect = sqrt(a_beta^2 * b_se^2  +  b_beta^2 * a_se^2  )
```

where
- `a_beta` and `a_se` are the total effect and SE from the step 1 MR analysis (exposure->mediator)
- `b_beta` and `b_se` are the total effect and SE from the step 2 MR analysis (mediator -> outcome), or the direct effect and SE from MVMR (mediator -> outcome)

*"Propagation of errors method"* – can be applied to both difference method and product method results

```
se.indirect = sqrt(se_total ^2 + se_direct^2)

se.indirect = sqrt(se_a^2 + se_b^2)
```

where
- `se_total` is the SE from the exposure -> outcome MR
- `se_direct` is the SE from exposure+mediator -> outcome MVMR (of exposure->outcome)

#### Mediation analysis full results

The indirect effect estimates calculated using Difference and Product methods were not consistent with one another in size and, in some cases, direction (Figure S6, Table S17). The confidence intervals based on SE calculated with Delta and PoE methods were also quite different, with PoE proving 10-fold magnitude SE, which resulted in wider confidence intervals. The inconsistent results made it difficult to draw any conclusions by evaluating both methods. Therefore, we carried out a simulation analysis (next section) to help us decide which combination of methods should be used for the mail analysis.

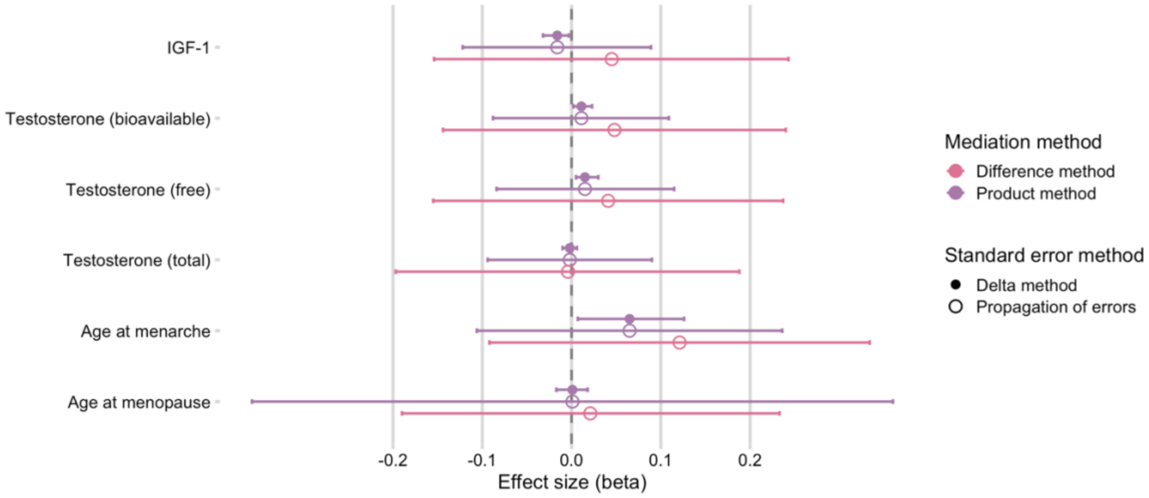


**Figure S6. Results of mediation analysis: indirect effect of childhood body size via mediators.** The point estimates represent the indirect effect (beta) that childhood body size has on breast cancer risk via the specified mediator (left-hand side labels). Indirect effects were calculated using Difference method (pink) and Product method (purple). The standard error of beta (which is used in confidence intervals calculation) was estimated using Delta method (filled circle) and "propagation of errors" (empty circle); both methods were applied to Product method's beta. Bars indicate 95% confidence intervals around the point estimates.

#### Simulation analysis for selecting the approach for mediation analysis

The available methods for mediation analysis produced results with some differences both in the estimation of indirect effect beta and standard error (described in the previous section). To estimate which combination of methods should be used in our study, we carried out a simulation analysis.

Firstly, based on the results from two-step and MVMR analyses we generated a model to simulate the individual-level data. This data was used to create simulated summary statistics and then apply the IVW method (linear regression) to estimate the effects from the simulated data (exposure -> mediator, mediator -> outcome, exposure + mediator -> outcome). Separate models were set up to represent each mediator individually. For each mediator, we carried out 1000 iterations of such simulations. In each iteration, we estimated indirect effect using Difference and Product methods from the simulated data and also calculated the corresponding SE using Delta and PoE methods.

Finally, we reviewed the distribution of indirect point estimates (from two mediation methods), i.e. we calculated standard deviation of the estimated betas across each iteration, and compared it with the average of the standard errors calculated by each method described above across all iterations, to assess the similarity across the pairs of methods. The results are available in Table S18.

Across all of the simulations there was a high level of consistency between the SD of beta from Product method and the mean of SE from Delta method (e.g. 0.0019 for IGF1). This similarity was observed for all mediators in the analysis. The other combination of methods (involving PoE method for SE estimation) produced much higher SE than the true value from the simulation. As a result, the combination of Product and Delta methods was chosen for the main analysis.
The simulation implementation is available here: https://github.com/mvab/simulation_for_MR_mediation

#### Proportion mediated calculation

The proportion of exposure effect mediated via a mediator is calculated as mediator indirect effect divided by the total effect beta [4]. For IGF-1, the proportion mediated was calculated as follows:

-0.016 / -0.411 = 0.039

(Product method-estimated indirect beta (Table S17) divided by the total effect beta (Table S1))

#### References


[1] S. Burgess, D. J. Thompson, J. M. B. Rees, F. R. Day, J. R. Perry, and K. K. Ong, “Dissecting causal pathways using mendelian randomization with summarized genetic data: Application to age at menarche and risk of breast cancer,” *Genetics*, vol. 207, no. 2, pp. 481–487, Oct. 2017.

[2] S. Burgess, R. M. Daniel, A. S. Butterworth, and S. G. Thompson, “Network Mendelian randomization: Using genetic variants as instrumental variables to investigate mediation in causal pathways,” *Int. J. Epidemiol.*, vol. 44, no. 2, pp. 484–495, 2015.

[3] C. L. Relton and G. Davey Smith, “Two-step epigenetic mendelian randomization: A strategy for establishing the causal role of epigenetic processes in pathways to disease,” *Int. J. Epidemiol.*, vol. 41, no. 1, pp. 161–176, 2012.

[4] A. R. Carter *et al.*, “Mendelian randomisation for mediation analysis: Current methods and challenges for implementation,” *European Journal of Epidemiology*. 2021.

[5] M. E. Sobel, “Asymptotic Confidence Intervals for Indirect Effects in Structural Equation Models,” *Sociol. Methodol.*, vol. 13, pp. 290–312, 1982.
